# Trends in fetal alcohol spectrum disorder (FASD) research: a systematic scoping review and bibliometric analysis

**DOI:** 10.1101/2024.04.29.24306557

**Authors:** Cheryl McQuire, Nessie Felicia Frennessen, James Parsonage, Molly Van der Heiden, David Troy, Luisa Zuccolo

**Author notes:** Corresponding author: Dr Cheryl McQuire,; Address: Bristol Medical School, Centre for Public Health, University of Bristol, Canynge Hall, 39 Whatley Road, Bristol, BS8 2PS, United Kingdom. Twitter/X: @cheryl_mcquire.

## Abstract

Fetal alcohol spectrum disorder (FASD) is a leading cause of neurodevelopmental disability globally. International health and policy organisations have highlighted an urgent need for improved prevention, diagnosis, and support. However, the evidence base needed to inform this is thought to be limited. We conducted a systematic scoping review and bibliometric analysis to describe trends in the volume and characteristics of original research on FASD published between 2000–2023 (Review 1). We compared the volume of published research on FASD to that of other neurodevelopmental disorders (Review 2). Searches were conducted in MEDLINE, Embase, CINAHL, PsychInfo (Review 1) and PubMed (Review 2). Eligible studies were original research articles with FASD terms in the title (Review 1) and all records with FASD, autism, or attention-deficit-hyperactivity disorder terms in the title (Review 2) published between 2000–2023. Data were summarised using descriptive statistics, narrative syntheses, and time-series plots to depict trends in common themes, countries of publication, sample characteristics, and research volume. This review found that FASD remains significantly under-researched. While there has been an increase in the number of original FASD research articles published annually over the last 23-years, this is significantly lower than what would be expected based on comparison with publication trends for other neurodevelopmental conditions, and the wider scientific literature. Further research is needed to understand the needs and impact of FASD across the lifespan and within different populations, to inform evidence-based prevention, policy, and support, and to advance progress in strength-based, stigma-reducing approaches to FASD research and practice.

**Statements and Declarations:** The authors declare no conflicts of interest

## Introduction

Fetal alcohol spectrum disorder (FASD) one of the most common preventable causes of lifelong neurodevelopmental disability [1]. Estimates suggest that FASD affects over 1% of children in the general population in 76 countries, amounting to 1700 infants being born with FASD every day, or 630,000 every year [2]. The European Region is understood to have the highest prevalence of FASD globally (2%) owing to high rates of prenatal alcohol use (Russia, United Kingdom [UK], Denmark, Belarus and Ireland have the highest estimated rates of PAE worldwide, ranging from 37 – 60%) [3,2].By way of comparison the estimated prevalence of autism and attention-deficit-hyperactivity-disorder (ADHD) within the European general population is 1% and 4%, respectively [4,5]. FASD rates are 10 – 40 times higher in ‘special subpopulations’ including Children in Care, correctional, special education, clinical and indigenous populations, compared to the general population [6]. Accordingly, international and national policy makers have called for urgent action to increase understanding, identification and support for those living with FASD [7–9].

Caused by prenatal alcohol exposure, FASD is characterised by significant neurodevelopmental problems including issues with learning, behaviour and adaptive functioning, and, in some cases, growth deficiency and a distinctive facial phenotype (present in approximately 10% of cases) [1]. Individuals with FASD are at an increased risk of adverse outcomes throughout the life course including poor physical and mental health, substance misuse, and contact with the criminal justice system [10–12]. Despite these far-reaching consequences, significant associated economic costs (e.g. $1.8 billion dollars in 2013 in Canada alone [13] and international evidence syntheses indicating that costs associated with FASD are 26% higher than costs for autism [14]) and unique opportunities for prevention, FASD is under-resourced and under-researched compared to other neurodevelopmental conditions [15–18].

There is substantial cross-country variation in the estimated prevalence of prenatal alcohol use and FASD [3,2]. However, the level of research and clinical provision for FASD within different populations does not appear to directly correlate with this level of need. For example, despite the UK having a higher estimated prevalence of prenatal alcohol exposure and FASD than countries such as the USA and Canada [2], and a significant research infrastructure, understanding and support for FASD in this country is limited [15,19]. Greater exploration of between-country differences in FASD research trends is warranted to gain insight into the nature and magnitude of such differences.

Examination of publication trends in FASD, including dominant areas of interest and research gaps is required to identify areas of progress and inform future research priorities. Comparison of FASD research trends with those of neurodevelopmental conditions, including autism and attention-deficit hyperactivity disorder (ADHD), is needed to contextualise observed trends and disparities. These comparisons will also provide a benchmark for understanding how FASD research compares to the general exponential increase in the volume of publications observed in the health literature in recent years [20].

Recent commentary, selected review, and ‘primer’ articles on FASD have suggested some priorities for future research and practice [21,17,22,18]. However, these articles have not been based on systematic searches and syntheses of the literature. Further empirical evidence on trends in FASD research, including analysis of between country and between-disorder differences in the volume and trends of outputs can provide important, novel insights into areas of progress and can identify gaps and likely future research directions, hence the importance of the present review. In this context, the objective of this systematic scoping review was to investigate trends in the volume and nature of research published on FASD between 2000 – 2023. We systematically searched and synthesised evidence on general trends in original FASD research (herein referred to as Review 1) and compared the volume of published articles on FASD with that of other neurodevelopmental disorders (Review 2). Specifically, we addressed the following questions:

### Review 1: Trends in original FASD Research

What is the amount, nature (study characteristics including: key themes, sample characteristics and country affiliated with the research team), and time trends of original FASD research published between 2000 - 2023?

### Review 2: Comparative review of volume and trends in FASD research and other neurodevelopmental disorders

How does the amount of research published on FASD overall, and over time, differ for FASD compared to other neurodevelopmental disorders, specifically autism and ADHD between 2000 - 2023?

Below we describe our general approach followed by specific methods and results for Review 1 and Review 2 in turn, followed by discussion and conclusion sections that consider findings from both reviews.

## Methods

### Methodological framework

This study was conducted in accordance with the JBI methodology for scoping reviews [23] and reported following the Preferred Reporting Items for Systematic reviews and Meta-Analyses extension for Scoping Reviews (PRISMA-ScR) [24]. We used a bibliometric analysis approach, which aims to ‘quantify a specific body of publications…to capture the state of knowledge in a specific field and contribute to insight regarding the underlying trends of a scientific discipline’[25]^(p.85)^.

### Protocol and registration

The pre-registered protocol for this review is available on the Open Science Framework platform via this link: https://osf.io/jyfpx/ [26].

### Ethical approval

Ethical approval was not required as this study used secondary data.

### Review 1: Trends in FASD Research

#### Eligibility criteria

Eligible studies were original research articles that included FASD or FASD subtype terms in the title (e.g. fetal alcohol syndrome [FAS], alcohol related neurodevelopmental disorder [ARND]), published between 2000 - 2023. Eligible study types included experimental and quasi-experimental study designs including randomized controlled trials, non-randomized controlled trials, before and after studies, and interrupted time-series studies. Analytical and descriptive observational studies including prospective and retrospective cohort studies, case-control studies and cross-sectional studies were also eligible, as were qualitative studies and original documentary-based and economic analyses, including modelling studies. We excluded studies of non-human animals, reviews, conference abstracts, commentaries, opinion pieces, expert consensus articles (including those reporting guideline development without novel data analysis or validation analyses), errata/corrections, letters, case studies (n=1) and pre-prints.

#### Search strategy

Preliminary searches of three electronic databases (MEDLINE, the Cochrane Database of Systematic Reviews, and JBI Evidence) in January 2023 revealed no protocols or published reviews looking at FASD research trends, indicating a need for the present review.

We searched the Cumulated Index to Nursing and Allied Health Literature (CINAHL), Embase, MEDLINE and PsychInfo for studies on FASD published in any language between 2000 – 2023. Initial searches were carried out between February – October 2023, with a final update search in February 2024. Search terms included a combination of Medical Subject Headings (MeSH) headings (e.g. *Fetal Alcohol Spectrum Disorders/*) and keywords corresponding to FASD and FASD subtypes, using wildcard operators to account for variations in spelling (e.g. f?etal alcohol spectrum disorder, f?etal alcohol syndrome, alcohol related neurodevelopmental disorder). The search strategy was developed in collaboration with University of Bristol library services and was based on strategies used in previous FASD review articles [27,28], identification of relevant keywords in preliminary MEDLINE and Embase searches and the review team’s existing knowledge of key publications.

Searches were limited to exclude studies of non-human animals. The MEDLINE search strategy is presented in Online Resource 1 (Table S1). This search was adapted, where necessary, to reflect search operators/processes in each bibliographic database.

#### Study screening and selection

All search records were imported into EndNote reference management software [29] and duplicates removed. Three members of the review team (CM, NFF and MvdH) screened studies for eligibility using a combination of software-assisted and manual screening. First, we identified records with FASD or FASD subtype terms in the title using the EndNote advanced search function, retaining these for further eligibility assessment.^a^ All other records were excluded. Duplicates were removed and we assessed the eligibility of remaining records based on the title, abstract, and full text, where necessary. Any disagreements for inclusion decisions were resolved through discussion between CM, NFF, MvdH, and/or with an additional reviewer (LZ).

#### Data Extraction

We (CM and DT) extracted data from eligible studies into a standardized Microsoft Excel spreadsheet [30]. The extracted data included: publication year, title, authors, journal, International Standard Serial Number (ISSN), author address and digital object identifier (DOI). We derived information about the country of origin for each publication using the ‘author address’ field of imported references, where available. For references that did not have an author address assigned, we (CM and DT) inferred the country affiliated with the research team by searching the name of the first/corresponding author to find their host institution. Where this information was not included, we inferred the country of origin from the study abstract, including details of where the study was conducted.

#### Data Analysis

We conducted a content analysis of themes covered in the FASD literature and between-country comparisons of the volume of outputs over time, using simple frequency counts and proportions. First, CM screened the titles, abstracts, and, where necessary, full texts to describe the main theme(s) of eligible studies (for example, ‘diagnosis/screening’, ‘intervention’). For studies including people with FASD, NFF and CM further categorised studies by age group, based on the terms/age groups reported in the title or abstract (e.g. children, adolescents, adults), and referring to the full text, if necessary. Studies were categorised by sample under one or more of the following groups: i) children with FASD (term ‘child(ren)’ in title, or aged 0 – 9 years according to abstract/full text); ii) adolescents with FASD (terms ‘youth(s)’, ‘adolescents’, or ‘young adults’ in title, or aged 10 – 19 [31] according to abstract/full text); iii) adults with FASD (aged 19+); iv) professionals (those working in a professional capacity with individuals with FASD [e.g. in education, health care, criminal justice, or policy settings]); v) caregivers (biological and non-biological, typically representing studies on the experiences and needs of those caring for someone with FASD); vi) birth mothers of people with FASD; vii) pregnant women; viii) postpartum women; ix) women of childbearing age (as defined by each study); x) biological fathers of people with FASD xi) other biological relations (e.g. siblings, grandparents of people with FASD); and so-called ‘special sub-populations’[6], including xii) looked-after-children (also known as children in care) and xiii) indigenous populations. Studies that did not include participants directly (e.g. documentary analyses) were classed as ‘other’, in terms of sample type.

It was possible for a study to be assigned to multiple thematic (e.g. ‘neurodevelopment’ and ‘diagnosis/screening’) or population subgroup categories (e.g. ‘children’ and ‘adolescents’), where applicable. Initial thematic and population-based groupings were performed in EndNote. Data on study characteristics, themes and sample were then imported from EndNote into Microsoft Excel, where they were screened a second time to verify initial thematic/subpopulation groupings, with amendments made where necessary (CM, LZ). Finally, extracted data were imported into R Studio software [32] to support further data analysis and visualisation using dplyr and ggplot2 packages [33,34].

## Results

Our searches identified 6,445 records for screening. After removing duplicates and applying our exclusion criteria, we identified 854 eligible original research studies on FASD, published between 2000 – 2023. Figure 1 provides a flow diagram of the selection process. The most common reasons for exclusion were that the record did not include a term for FASD or one of the subtypes in its title (42% of identified records), or that it was not original research (e.g. commentary or review article [28%]). Below we describe trends in FASD research by study theme, country of publication, and population of interest. Online Resource 2 provides an output table of all eligible studies. This can be filtered by theme (e.g. ‘neurodevelopment’) and includes details of study characteristics (sample population, country of corresponding/lead author, year of publication, authors, title, journal and DOI, where reported).^b^

**Fig. 1.**
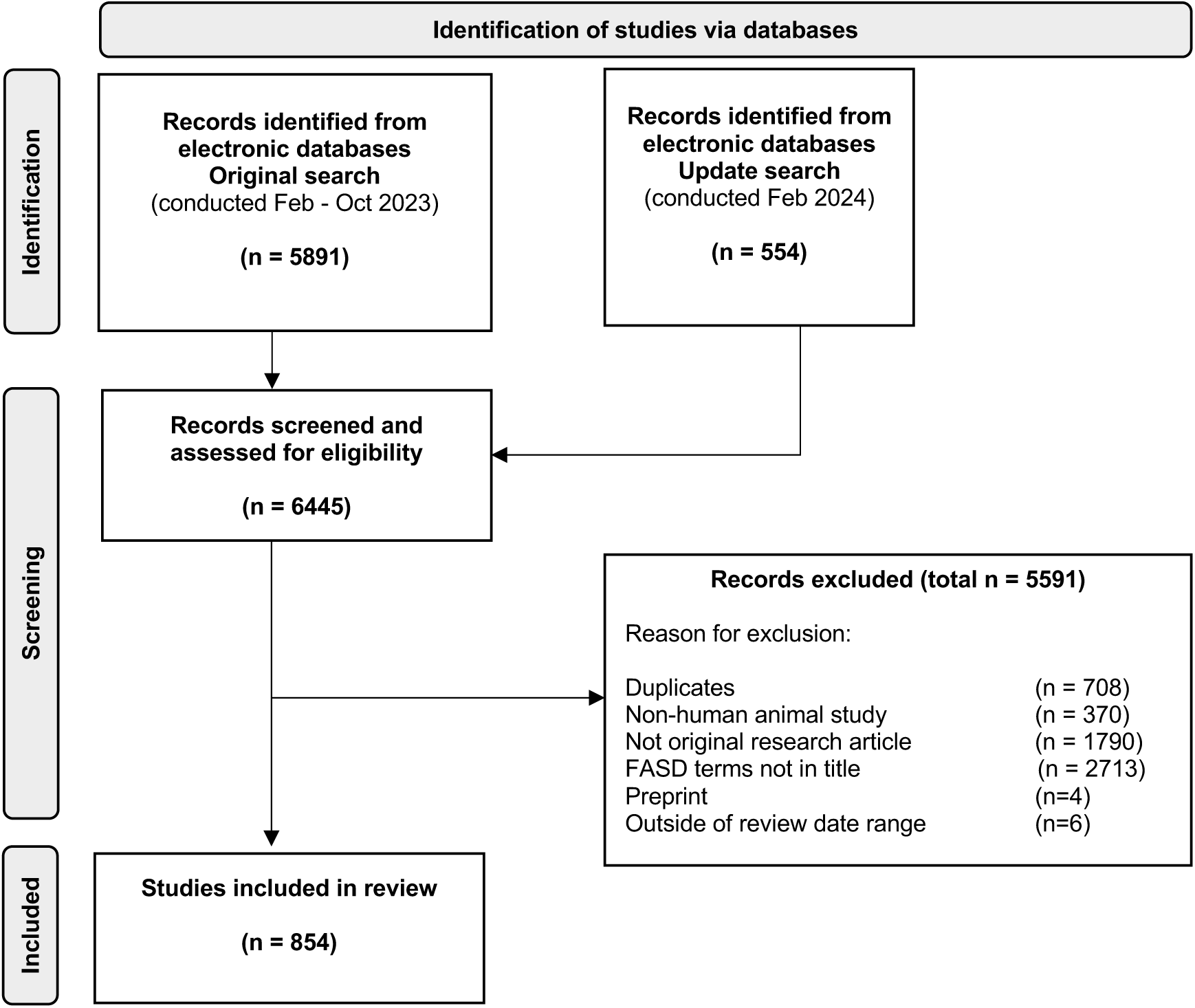
PRISMA flow chart depicting study identification and selection process [35]

### Overall time trend in FASD research publications

The number of original research articles published annually on FASD increased over time (2000 – 2023) but remained relatively low in absolute terms Figure 2. In the results section for Review 2 we will assess how trends in the volume of research published on FASD compares to that of other neurodevelopmental disorders (autism and ADHD). In this section, we present results by country of origin, article theme and sample population.

**Fig. 2.**
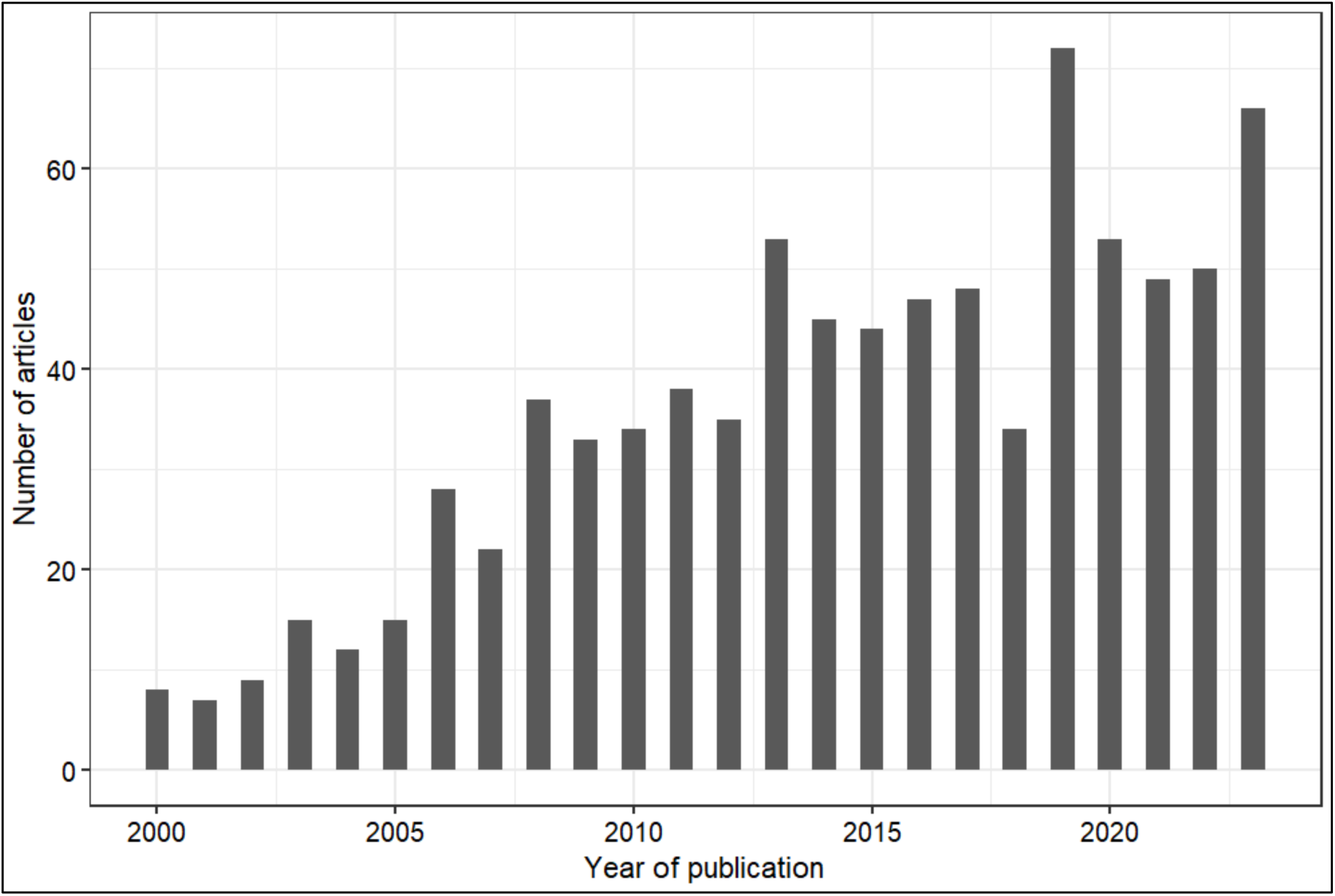
Number of original research articles with FASD terms in the title published per year between 2000 – 2023

### FASD research publications by country of origin

Eligible publications originated from 31 countries, however most of these countries (68%) had fewer than 10 studies published on FASD over the entire 23-year review period (Figure 3). Nearly two thirds (557/854 [65%]) of all eligible articles were produced by research teams from North America. The five countries that produced the most FASD research articles were the United States of America (38% of eligible articles), Canada (27%), Australia (8%), South Africa (6%) and the United Kingdom (4%). Figure 4 presents time trends in the number of publications per year for each of these five countries. We can see that authors from the USA dominate research published on FASD from the year 2000 onwards, with notable increases in the volume of research published by teams from other countries from 2005, most notably Canada, followed by Australia and the UK.

**Fig. 3.**
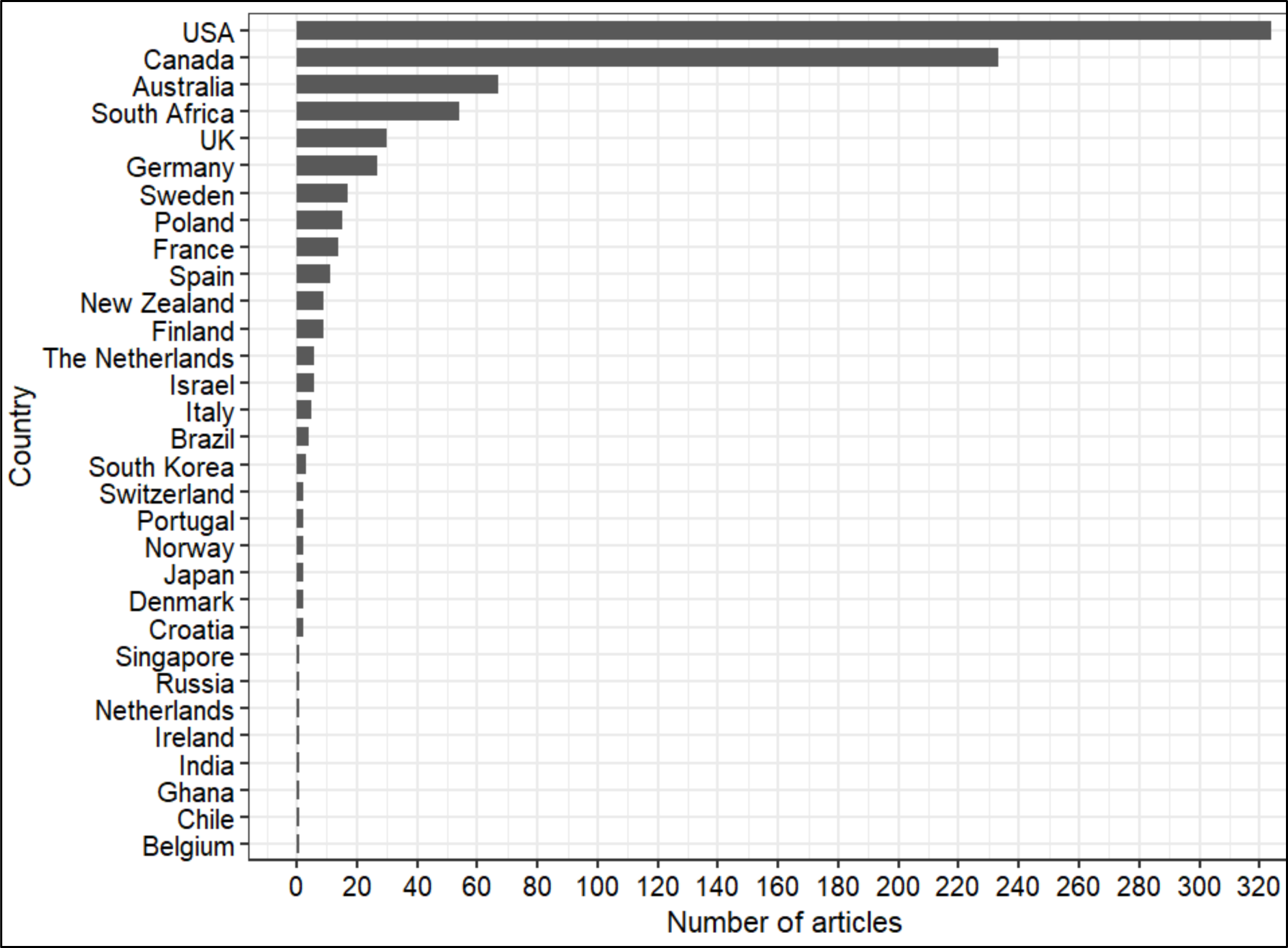
Number of eligible articles on FASD by country of the corresponding or lead author

**Fig. 4.**
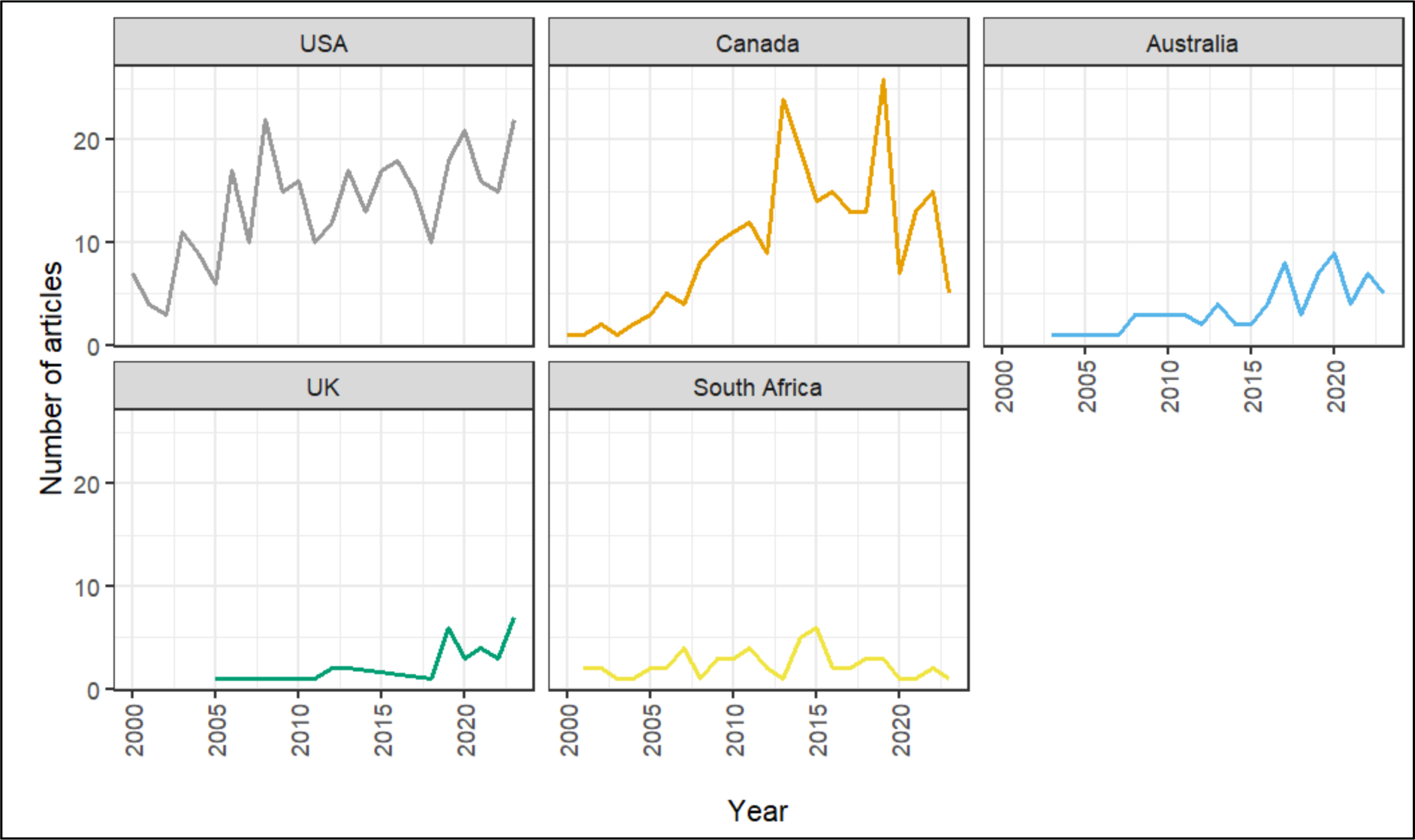
Number of eligible articles on FASD over time by the five most common countries of origin

### FASD research article themes

We derived 29 common themes to describe the primary focus of eligible FASD articles (Figure 5). Research on the neurodevelopmental phenotype of FASD (e.g. learning, memory, attention, executive functioning) was the most common theme (234/854 [27%] of eligible studies), followed by the closely related themes of diagnosis/screening for FASD (166/854 [19%]), neuroscience (e.g. brain imaging; 99/854 [12]), and research on FASD interventions (100/854 [12%]). Online Resource 1 (Figure S2) presents a summary of time-trends for all of the themes that had at least 50 eligible articles published within the study period.

**Fig. 5.**
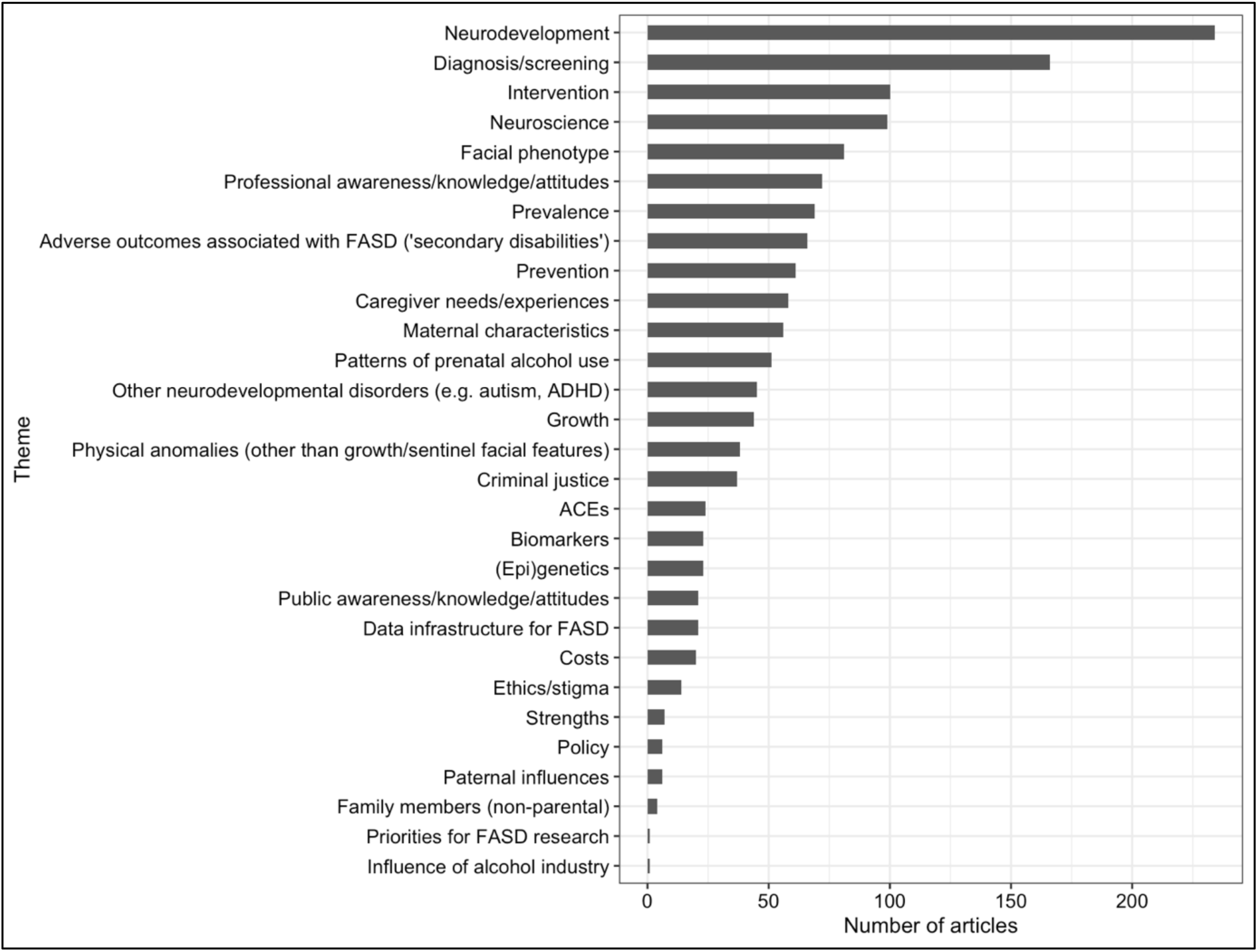
Number of articles for each thematic grouping, published between 2000 – 2023

Next, we explored time trends in the volume of research published for each component of the FASD phenotype – neurodevelopmental functioning, growth, and facial phenotype (Figure 6). Here, we can see a marked increase in research on the neurodevelopmental phenotype from 2005 onwards.

**Fig. 6.**
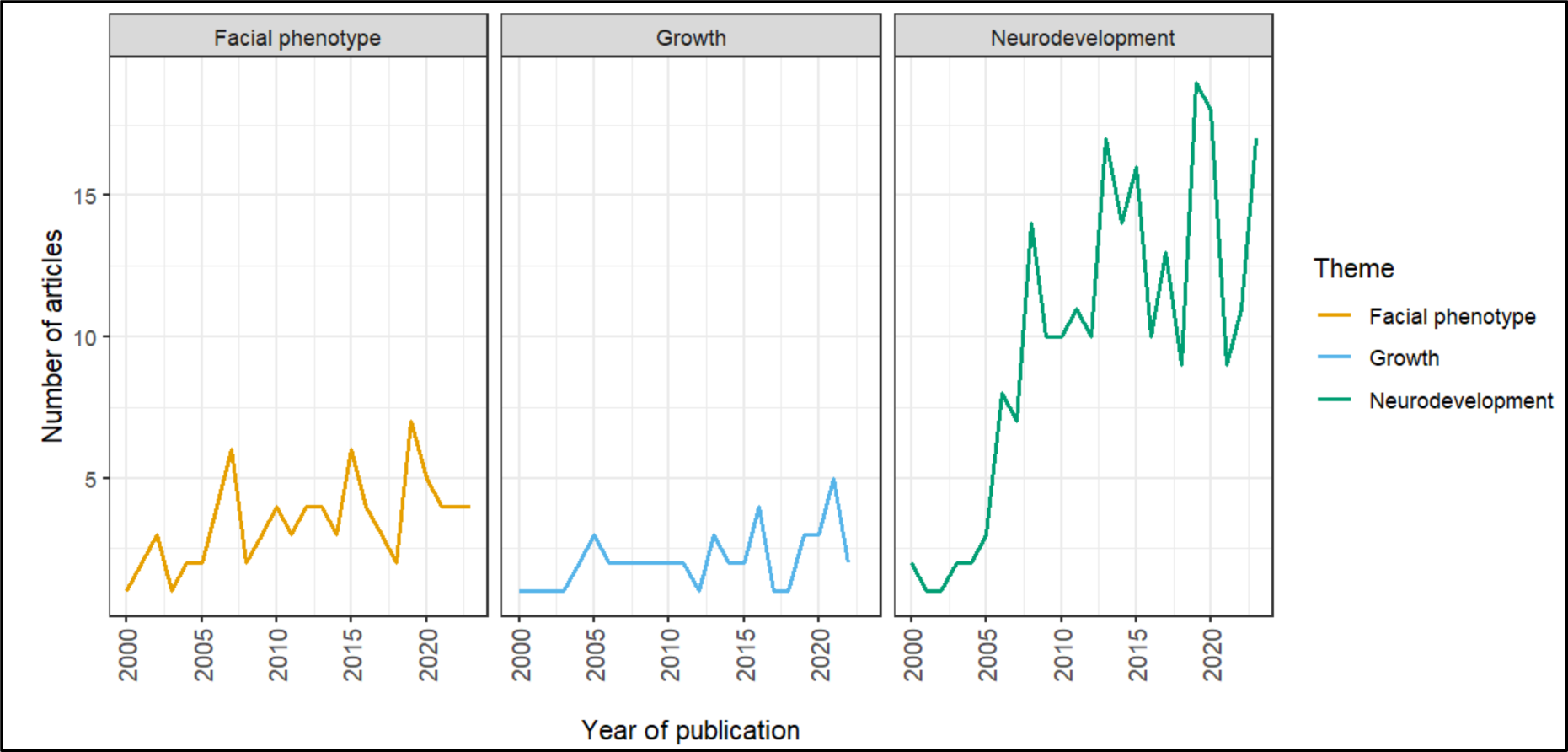
Number of articles published annually on the three core aspects of the FASD phenotype

### FASD research publications by sample population

Finally, we categorised studies by sample population (Figure 7). There was limited research that had a primary focus on biological fathers (1% of eligible studies), siblings (0.4%), and grandparents of children with FASD, compared to the percentage of eligible studies that included women of reproductive age or birth mothers of children with FASD (15% in total, consisting of birth mothers [9%], pregnant/postpartum women [4%], women of childbearing age [2%]). Children with FASD featured in 59% of eligible studies, compared to 23% and 14% of studies with adolescents and adults, respectively.

**Fig. 7.**
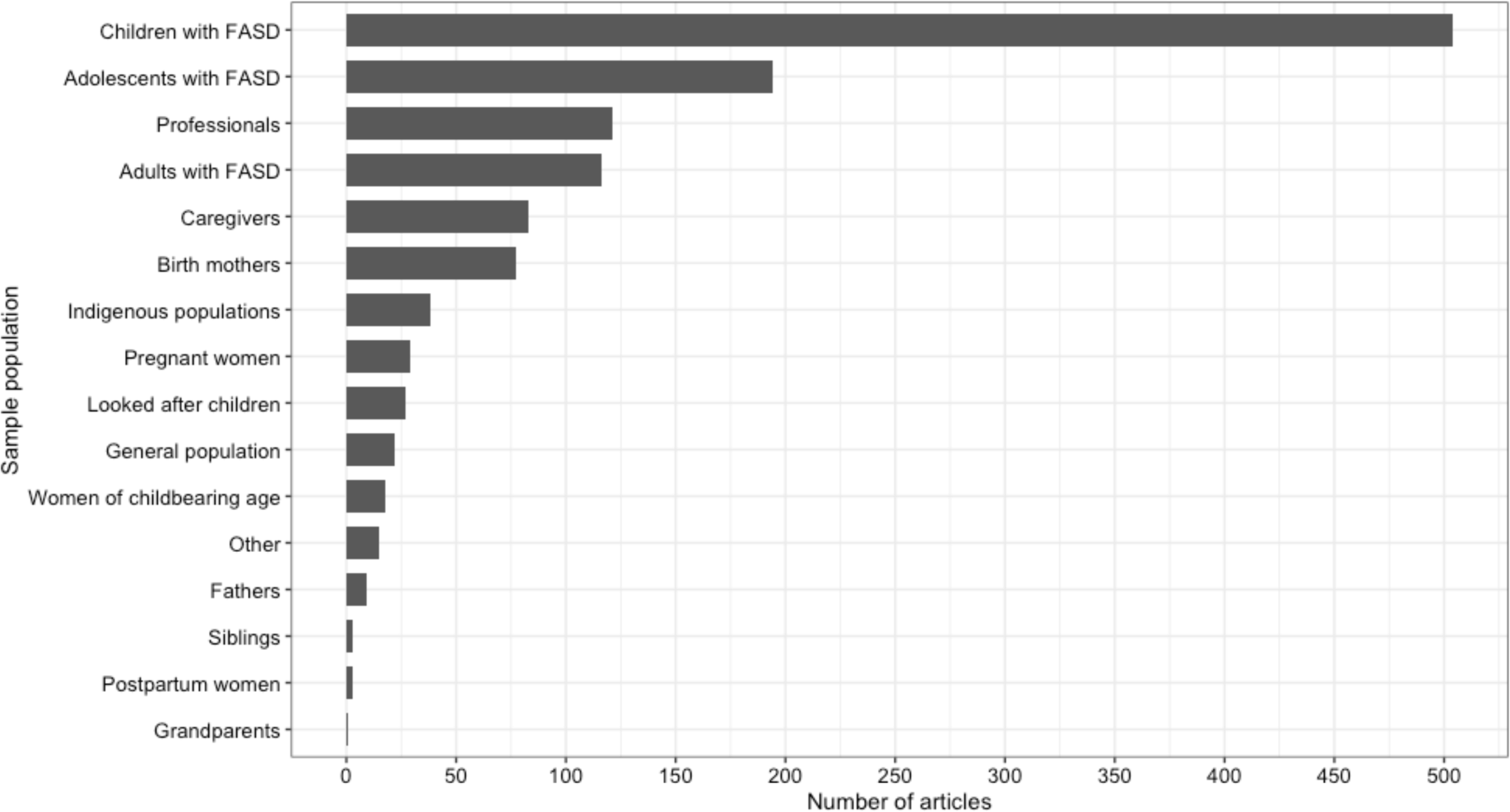
Number of articles published by sample population

We also looked at trends in FASD research sample groups featuring children, adolescents, and adults (Figure 8). While the research output around adolescents and adults has increased over time, research into children with FASD has remained dominant.

**Fig. 8.**
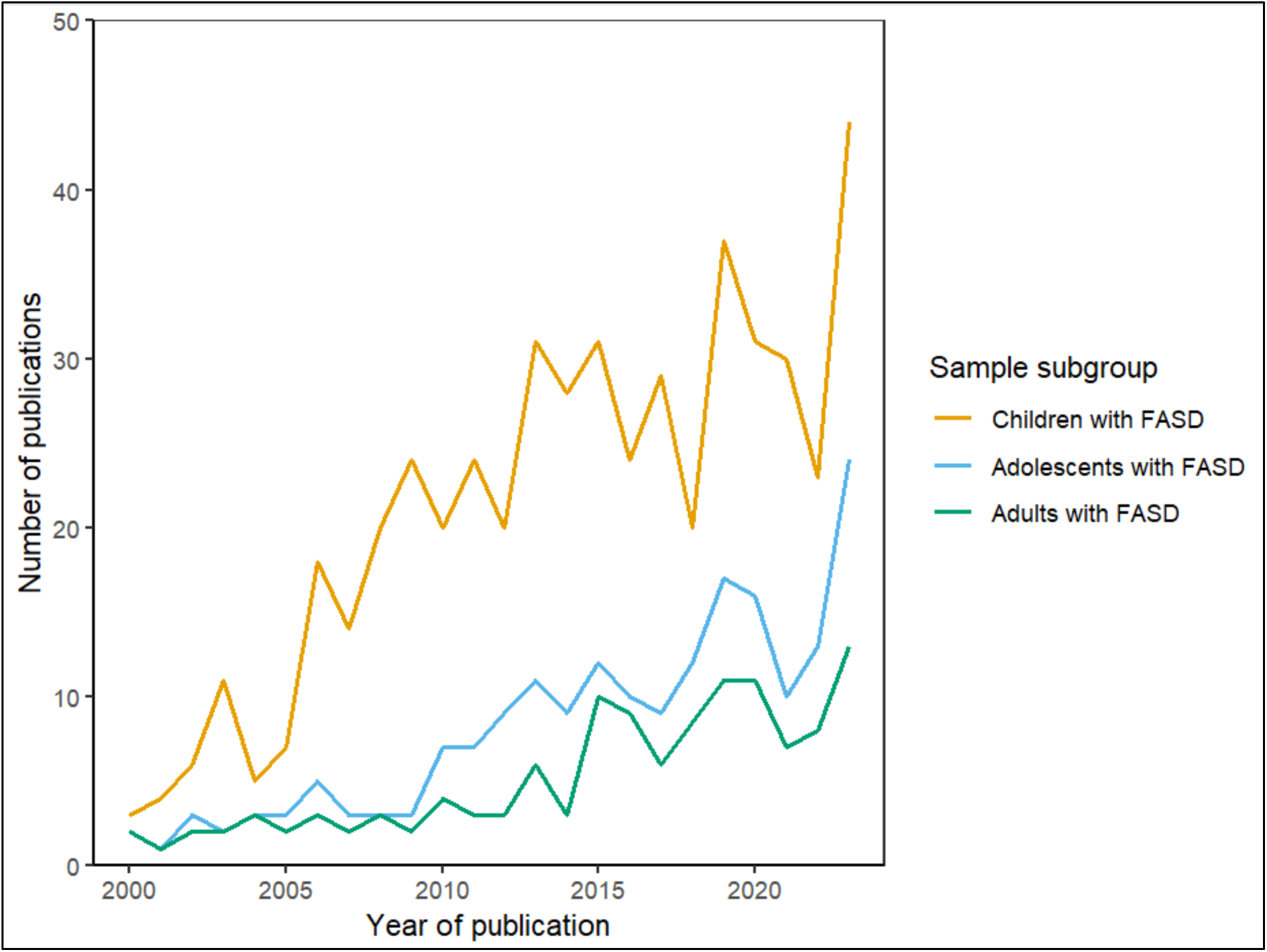
Number of articles published annually with children, adolescents, and/or adults with FASD as sample participants

### Review 2: Comparative review of the volume of FASD research, compared to other neurodevelopmental disorders

The aim of this review was to compare time trends in FASD research publications to those for other neurodevelopmental disorders, specifically ADHD and autism.

#### Eligibility criteria

All records published between 2000 – 2023 with terms relating to ‘autism’, ‘fetal alcohol spectrum disorder’ or ‘ADHD’ in the title were eligible for inclusion. Studies of non-human animals were excluded.

#### Search strategy

We searched PubMed for records with terms relating to FASD, ADHD or autism in the title (Supplement 1, Table S3). We filtered results to exclude non-human animal studies and to limit results to records published between 2000 – 2023.

#### Study screening and selection

We included all records identified in our PubMed search strategy.

#### Data Extraction and analysis

We extracted the number of results by year for FASD, autism and ADHD into Microsoft Excel, using the PubMed ‘download CSV’ function under ‘Results by Year’.

#### Data Analysis

We used Microsoft Excel to produce descriptive statistics and comparative time series plots of the FASD, autism and ADHD publications (Figure 9). Our searches retrieved a total of 64,069 records that had terms for any of these neurodevelopmental conditions in the title (FASD, autism, or ADHD). Of this combined total, the majority (60%) of records were for autism, followed by ADHD (38%). Only 2% of records fell under the FASD category.

**Fig. 9.**
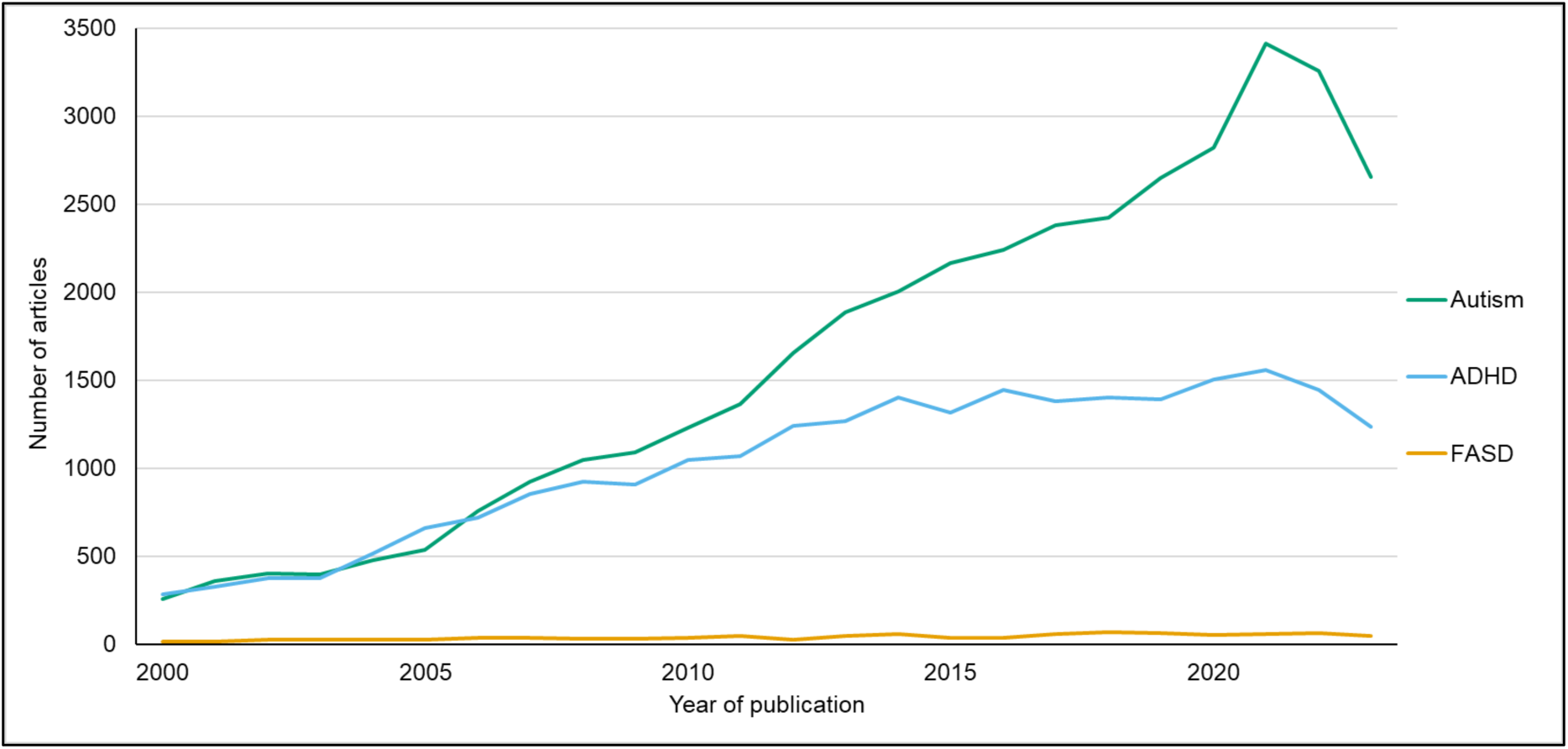
Number of PubMed records published annually with FASD, ADHD, or autism terms in the title

## Discussion

### Key findings

While there has been an increase in the volume of original FASD research articles published annually between the years 2000 – 2023, our review indicates that this trend is significantly lower than expected, based on the exponential increase, or ‘deluge’, of scientific outputs observed in many fields [20]. Our searches indicated that FASD publications made up just 2% of the literature published on some of the most common forms of neurodevelopmental conditions during this time (autism, ADHD and FASD [4,36,5,2]). There were marked differences in the amount of research published by country, with approximately two-thirds (65%) of all FASD outputs led by teams in North America (USA and Canada). Our analysis of research themes shows a relative focus on diagnosis and screening, compared to prevention and intervention. Overall, research related to the neurodevelopmental phenotype of FASD was the most common, representing over a quarter (27%) of eligible studies.

Exploration of time trends related to the different components of the FASD phenotype – sentinel facial features (smooth philtrum, thin upper lip, short palpebral fissure length), growth restriction and neurodevelopmental abnormalities (e.g. learning, memory, attention)[1] – revealed a change from research that focused mainly on physical indicators (especially the facial phenotype) up until 2005, towards studies focusing on the neurodevelopmental phenotype in more recent years. This is likely to reflect an evolution in conceptualisations of FASD – beginning with fetal alcohol syndrome (FAS), a term coined by dysmorphologists in the late 1960’s/early 1970s to describe (predominantly) the facial and growth anomalies caused by prenatal alcohol exposure [37,38], followed by guidelines with a much greater focus on neurodevelopmental aspects of FASD from the late 1990’s onwards [39–48,9]. Despite broad consensus on the neurodevelopmental aspects of FASD, diagnostic guidelines remain heterogenous in terms of the specific criteria and thresholds used to indicate neurodevelopmental impairment. This complexity, coupled with a search for a definitive neurodevelopmental phenotype for FASD are likely to have contributed to the relative proliferation of research on this theme. It is now known that only ∼10% of individuals with FASD present with sentinel facial features, thus FASD is largely regarded as an ‘invisible disability’ [1]. Nevertheless, sentinel facial features remain an important aspect in recognising FASD, particularly as a distinct neurodevelopmental profile for FASD remains elusive.[49–52]

### Identification of evidence gaps

Overall, research on FASD remains limited to a minority of countries and communities internationally. Publications on FASD were attributed to 31 countries, however most of these countries (52%) had fewer than five articles on FASD across the entire 23-year review period. Furthermore, cross-country variation in the number of research outputs, was not consistent with country-level research capacity (based on research spend and workforce [53,54]), or the population prevalence of FASD [2]. For example, despite having a world-leading research infrastructure, and the fourth highest prevalence of prenatal alcohol exposure globally [3], only 4% of eligible studies originated from the UK.

Since FASD is a lifelong condition, the relative scarcity of research involving adolescents and adults with FASD represents an important evidence gap.[1] Available evidence on adolescents and adults with FASD point to challenges with physical and mental health, and in daily living, particularly during the transition from child to adult services [10,11]. While some transition planning tools are available, research evaluating the acceptability and effectiveness of these tools, and of ongoing functional and needs-based service provision to support successful transition to adulthood (e.g. independent living) remains scarce [55].

We found limited research on biological fathers, siblings, and grandparents of children with FASD. This is notable since preliminary evidence indicates that prenatal alcohol exposure and FASD risk are influenced by social-psychological and biological factors [27,56]. For example, relative to controls, mothers of children with FASD are more likely to report using alcohol to cope with childhood trauma and interpersonal violence and to have experienced coercion from a partner to consume alcohol in pregnancy [56,27,57]. Biological fathers and grandparents of children with FASD are more likely to have abused alcohol, exhibit a risk genotype for heavy alcohol use, may pass on the risk of heavy alcohol use to subsequent generations through epigenetic mechanisms, and could provide a model of problematic alcohol use that could be acquired by children through social learning [58,56,27,59,60].

Stigma has been highlighted as a multi-level issue that negatively impacts FASD research and care [17]. Despite these issues, our review highlighted a paucity of stigma and strength-based research on FASD. Narratives that suggest an inevitability of negative outcomes among people living with FASD, coupled with a neglect of strength-based approaches, can lead to marginalisation and exclusion from services and poorer long-term outcomes [61–63]. Stigma leading to feelings of guilt or shame, experienced by women or parents of children with FASD may prevent or delay them from seeking support, and cultural stigmatisation, including perceptions of FASD as an ‘indigenous issue’, has also been reported [17]. Our finding that FASD research remains focused on maternal contributions to FASD (with limited evidence on paternal and wider familial/social influences) is likely to further compound the stigma, shame, and blame that is targeted towards women and mothers [64] and unduly narrow the focus of attempts to fully understand and address wider risk factors for PAE and

FASD. Developing approaches to reduce stigma are crucial to the development of successful initiatives to reduce and prevent prenatal alcohol exposure, and for improving access to support for those who are affected by prenatal alcohol use, including families and children [17].

### Strengths and limitations

This is the first review to systematically map the landscape of FASD research over time in order to understand trends by study topic, participant characteristics, country of publication and corresponding research gaps (Review 1). Comparing trends in FASD research with that for other neurodevelopmental conditions adds important context to these observed trends (Review 2). Our methodological approach followed models of best practice and included comprehensive searches across five major bibliographic databases.

Since this was a systematic scoping review, the primary aim of Review 1 was to identify and characterise the breadth of evidence available on FASD [65] as opposed to a full systematic review, which aims to collate and appraise all empirical evidence that fits pre-specified eligibility criteria (including supplementary searches of study reference lists, for example) [66]. Therefore, it is possible that some eligible studies were missed. Nevertheless, we contend that the scoping review methodology was most appropriate to our aim of characterising trends in original FASD research in human populations over the last 23 years.

One limitation of our comparative review (Review 2; exploring trends in FASD outputs with those for ADHD and autism) was that - owing to the large number of records identified across these neurodevelopmental conditions (∼65,000) and limited study resource - it was not possible to carry out a manual sift in order to limit results to original research studies with a distinct focus on one disorder (either FASD, autism, or ADHD). This is likely to have resulted in a high sensitivity to capture records with these terms in the title, but a lower specificity for retaining only relevant records for each of these neurodevelopmental conditions. It is also important to note that due to high rates of co-morbidity and misdiagnosis between FASD, ADHD and autism, these disorders, and the corresponding literature, are not mutually exclusive [67,68,52]. Therefore, comparisons of the literature base for each of these conditions, as discrete entities, is likely to be a simplification.

Categorisation of studies by theme, country of origin and participant group was performed by several members of the review team and included verification checks (double coding) across the review process. Nevertheless, there is likely to have been an element of subjectivity, particularly in the thematic coding (which relied on researchers’ assessment of the primary focus of each article), and in participant subgroup categorisation, which – for reasons of efficiency - relied predominantly on terms reported in the title and/or abstract. This is likely to be an approximation as our exploration of some full text articles revealed that this may not reflect the full composition of the sample (e.g. some studies that reported ‘children’ in the title had age ranges up to young adulthood when the full text was examined). Finally, given the low absolute number of eligible studies for FASD over the review period it must be noted that depiction and interpretation of time-trends, particularly within, for example, less common thematic groupings, should be considered illustrative only, especially since there appears to be significantly variability in publication volume over time within many of these groupings.

## Conclusion

FASD remains significantly under-studied. Further research is needed to understand the needs and impact of FASD within different populations across the lifespan, to support development of effective strategies for prevention and intervention, and to advance progress in strength-based, stigma-reducing approaches to FASD research and practice.

## Supporting information

Supplementary information 1_search strategies and supplementary figure

Supplementary information 2_Searchable output tables by theme

## Data Availability

Review article using secondary data (publicly available)

## Abbreviations

ADHD: Attention deficit hyperactivity disorder
ARBD: Alcohol related birth defect
ARND: Alcohol related neurodevelopmental disorder
CINAHL: Cumulated Index to Nursing and Allied Health Literature
ISSN: International Standard Serial Number
FASD: Fetal alcohol spectrum disorder
FAS: Fetal alcohol syndrome
PAE: Prenatal alcohol exposure
pFAS: Partial fetal alcohol syndrome
PRISMA-ScR: Preferred Reporting Items for Systematic reviews and Meta-Analyses extension for Scoping Reviews
MeSH: Medical Subject Headings

## Acknowledgments

This project was supported in part by the National Institute for Health Research (NIHR) School for Public Health Research (SPHR) (Grant Reference Number PD-SPH-2015-10025, Postdoctoral Launching Fellowship awarded to CM; and Grant Reference Number NIHR 204000, Pre-Doctoral Fellowship awarded to JP). The views expressed are those of the author(s) and not necessarily those of the NIHR or the Department of Health and Social Care; Grant MR/N0137941/1 for the GW4 BIOMED MRC DTP, awarded to the Universities of Bath, Bristol, Cardiff and Exeter from the Medical Research Council (MRC)/UKRI (awarded to NFF). LZ was supported by core funding by the Human Technopole, Health Data Science Centre.

## Declarations

### Author contributions

CM conceived of the study, developed the protocol, and search strategies, ran database searches, developed the extraction template, performed data extraction and analyses, and produced the initial draft of the manuscript; NFF contributed to data extraction and analysis, including grouping studies by sample population; JP contributed to the analysis, including graphical and tabular presentation of findings, and drafting of the initial manuscript; DT contributed to data extraction, including allocation of country of origin to research articles; MvdH contributed to the development and execution of search strategies; LZ developed the initial idea for this study with CM, contributed to inclusion decisions and categorisations, and provided input on all aspects of the study. All authors have contributed and agreed to the content of the submitted manuscript.

### Ethical approval

Ethical approval was not required as this study used secondary data.

## Footnotes

a Search terms for FASD and FASD subtypes for EndNote screening were: ‘fetal alcohol spectrum disorder’, ‘foetal alcohol spectrum disorder’, ‘FASD’, ‘FAS’, ‘fetal alcohol syndrome’, ‘foetal alcohol syndrome’, ‘alcohol related neurodevelopmental disorder’, ‘ARND’, ‘pFAS’, ‘ARBD’, ‘alcohol-related birth defect’

b Note that study information is taken directly from the record supplied by the bibliographic database.

## References

1. Popova, S., Charness, M. E., Burd, L., Crawford, A., Hoyme, H. E., Mukherjee, R. A. S., et al. (2023). Fetal alcohol spectrum disorders. Nature Reviews Disease Primers, 9(1), 11. 10.1038/s41572-023-00420-x

2. Lange, S., Probst, C., Gmel, G., Rehm, J., Burd, L., & Popova, S. (2017). Global prevalence of fetal alcohol spectrum disorder among children and youth: a systematic review and meta-analysis. JAMA Pediatrics, 171(10), 948–956. 10.1001/jamapediatrics.2017.1919

3. Popova, S., Lange, S., Probst, C., Gmel, G., & Rehm, J. (2017). Estimation of national, regional, and global prevalence of alcohol use during pregnancy and fetal alcohol syndrome: a systematic review and meta-analysis. Lancet Glob Health, 5(3), e290–e299. 10.1016/s2214-109x(17)30021-9

4. Zeidan, J., Fombonne, E., Scorah, J., Ibrahim, A., Durkin, M. S., Saxena, S., et al. (2022). Global prevalence of autism: A systematic review update. Autism Res, 15(5), 778–790. 10.1002/aur.2696

5. Polanczyk, G., de Lima, M. S., Horta, B. L., Biederman, J., & Rohde, L. A. (2007). The worldwide prevalence of ADHD: a systematic review and metaregression analysis. Am J Psychiatry, 164(6), 942–948. 10.1176/ajp.2007.164.6.942

6. Popova, S., Lange, S., Shield, K., Burd, L., & Rehm, J. (2019). Prevalence of fetal alcohol spectrum disorder among special subpopulations: a systematic review and meta-analysis. Addiction, 114(7), 1150–1172. 10.1111/add.14598

7. Department of Health and Social Care (DHSC) (2021). Fetal alcohol spectrum disorder: health needs assessment. DHSC, London. https://www.gov.uk/government/publications/fetal-alcohol-spectrum-disorder-health-needs-assessment/fetal-alcohol-spectrum-disorder-health-needs-assessment. Accessed 25 April 2024.

8. Jonsson, E., Salmon, A., & Warren, K. R. (2014). The International Charter on Prevention of Fetal Alcohol Spectrum Disorder. Lancet Global Health, 2(3), 135–137. 10.1016/S2214-109X(13)70173-6

9. National Institute for Health and Care Excellence (NICE) (2022). Fetal alcohol spectrum disorder quality standard. NICE, London. https://www.nice.org.uk/guidance/qs204. Accessed 25 April 2024.

10. McLachlan, K., Flannigan, K., Temple, V., Unsworth, K., & Cook, J. L. (2020). Difficulties in daily living experienced by adolescents, transition-aged youth, and adults with fetal alcohol spectrum disorder. alcoholism, clinical and experimental research, 44(8), 1609–1624. 10.1111/acer.14385

11. Popova, S., Temple, V., Dozet, D., O’Hanlon, G., Toews, C., & Rehm, J. (2021). Health, social and legal outcomes of individuals with diagnosed or at risk for fetal alcohol spectrum disorder: Canadian example. Drug and alcohol dependence, 219, 108487. 10.1016/j.drugalcdep.2020.108487

12. Streissguth, A. P., Bookstein, F. L., Barr, H. M., Sampson, P. D., O’Malley, K., & Young, J. K. (2004). Risk factors for adverse life outcomes in fetal alcohol syndrome and fetal alcohol effects. Journal of Developmental & Behavioral Pediatrics, 25(4), 228–238. 10.1097/00004703-200408000-00002

13. Popova, S., Lange, S., Burd, L., & Rehm, J. (2016). The Economic Burden of Fetal Alcohol Spectrum Disorder in Canada in 2013. Alcohol and alcoholism, 51(3), 367–375. 10.1093/alcalc/agv117

14. Greenmyer, J. R., Klug, M. G., Kambeitz, C., Popova, S., & Burd, L. (2018). A multicountry updated assessment of the economic impact of fetal alcohol spectrum disorder: costs for children and adults. Journal of Addiction Medicine, 12(6). 10.1097/ADM.0000000000000438

15. Scholin, L., Mukherjee, R. A. S., Aiton, N., Blackburn, C., Brown, S., Flemming, K. M., et al. (2021). Fetal alcohol spectrum disorders: an overview of current evidence and activities in the UK. Archives of disease in childhood, 106(7), 636–640. 10.1136/archdischild-2020-320435

16. Popova, S., Dozet, D., Temple, V., McFarlane, A., Cook, J., & Burd, L. (2024). Fetal alcohol spectrum disorder diagnostic clinic capacity in Canadian Provinces and territories. PLoS One, 19(4), e0301615. 10.1371/journal.pone.0301615

17. Flannigan, K., Pei, J., McLachlan, K., Harding, K., Mela, M., Cook, J., et al. (2021). Responding to the unique complexities of fetal alcohol spectrum disorder. Front Psychol, 12, 778471. 10.3389/fpsyg.2021.778471

18. Burd, L., & Popova, S. (2019). Fetal alcohol spectrum disorders: fixing our aim to aim for the fix. Int J Environ Res Public Health, 16(20). 10.3390/ijerph16203978

19. Aiton, N. (2021). Neglect of fetal alcohol spectrum disorder must end. BMJ, 375, n2969. 10.1136/bmj.n2969.

20. Chu, J. S. G., & Evans, J. A. (2021). Slowed canonical progress in large fields of science. Proceedings of the National Academy of Sciences, 118(41), e2021636118. 10.1073/pnas.2021636118

21. Popova, S., Charness, M. E., Burd, L., Crawford, A., Hoyme, H. E., Mukherjee, R. A. S., et al. (2023). Fetal alcohol spectrum disorders. Nat Rev Dis Primers, 9(1), 11. 10.1038/s41572-023-00420-x

22. Popova, S., Dozet, D., & Burd, L. (2020). Fetal alcohol spectrum disorder: can we change the future? Alcohol: Clinical and Experimental Research, 44(4), 815–819. 10.1111/acer.14317

23. Peters MDJ, G. C., McInerney P, Munn Z, Tricco AC, Khalil, H., (2020). Chapter 11: Scoping Reviews. In M. Z. Aromataris E (Ed.), JBI Manual for Evidence Synthesis. https://jbi-global-wiki.refined.site/space/MANUAL/4687342/Chapter+11%3A+Scoping+reviews. Accessed 25 April 2024.

24. Tricco, A. C., Lillie, E., Zarin, W., O’Brien, K. K., Colquhoun, H., Levac, D., et al. (2018). PRISMA extension for scoping reviews (PRISMA-ScR): checklist and explanation. Annals of internal medicine, 169(7), 467–473. 10.7326/M18-0850

25. Soares, E. E., Thrall, J. N., Stephens, T. N., Rodriguez Biglieri, R., Consoli, A. J., & Bunge, E. L. (2020). Publication trends in psychotherapy: bibliometric analysis of the past 5 decades. Am J Psychother, 73(3), 85–94, 10.1176/appi.psychotherapy.20190045

26. McQuire, C., van der Heiden, M., Troy, D., & Zuccolo, L. (2023). Trends in fetal alcohol spectrum disorder (FASD) research: protocol for a systematic scoping review and bibliometric analysis. https://osf.io/jyfpx/. Accessed 25 April 2024.

27. McQuire, C., Daniel, R., Hurt, L., Kemp, A., & Paranjothy, S. (2020). The causal web of foetal alcohol spectrum disorders: a review and causal diagram. Eur Child Adolesc Psychiatry, 29(5), 575–594. 10.1007/s00787-018-1264-3

28. Esper, L. H., & Furtado, E. F. (2014). Identifying maternal risk factors associated with fetal alcohol spectrum disorders: a systematic review. European Child and Adolescent Psychiatry, 23(10), 877–889, 10.1007/s00787-014-0603-2

29. The EndNote Team (2013). EndNote. (EndNote 20 ed.). Philadelphia, PA: Clarivate.

30. Microsoft Corporation (2024). Microsoft Excel for Mac. (Version 16.84 ed.).

31. Sacks, D., Society, C. P., & Committee, A. H. (2003). Age limits and adolescents. Paediatr Child Health, 8(9), 577–578. 10.1093/pch/8.9.577

32. R Core Team (2023). R: A language and environment for statistical computing. (Version 1.4.1103 ed.). Vienna, Austria: R Foundation for Statistical Computing.

33. Wickham H, F. R., Henry L, Müller K, (2022). dplyr: A Grammar of Data Manipulation. https://dplyr.tidyverse.org, https://github.com/tidyverse/dplyr. Accessed 25 April 2024.

34. Wickham H (2016). ggplot2: Elegant Graphics for Data Analysis. Springer-Verlag New York. https://ggplot2.tidyverse.org. Accessed 25 April 2024.

35. Page, M. J., McKenzie, J. E., Bossuyt, P. M., Boutron, I., Hoffmann, T. C., Mulrow, C. D., et al. (2021). The PRISMA 2020 statement: an updated guideline for reporting systematic reviews. BMJ, 372, n71. 10.1136/bmj.n71

36. Salari, N., Rasoulpoor, S., Rasoulpoor, S., Shohaimi, S., Jafarpour, S., Abdoli, N., et al. (2022). The global prevalence of autism spectrum disorder: a comprehensive systematic review and meta-analysis. Italian Journal of Pediatrics, 48(1), 112, 10.1186/s13052-022-01310-w

37. Lemoine, P., Harousseau, H., Borteyru, J., & Menuet, J. (2003). Children of alcoholic parents— observed anomalies: discussion of 127 cases. Therapeutic drug monitoring, 25(2), 132–136. 10.1097/00007691-200304000-00002

38. Jones, K., Smith, D., Ulleland, C., & Streissguth, A. (1973). Pattern of malformation in offspring of chronic alcoholic mothers. The Lancet, 301(7815), 1267–1271. 10.1016/s0140-6736(73)91291-9

39. Astley, S. J., & Clarren, S. K. (2000). Diagnosing the full spectrum of fetal alcohol-exposed individuals: introducing the 4-digit diagnostic code. Alcohol and alcoholism, 35(4), 400–410. 10.1093/alcalc/35.4.400

40. Hoyme, H. E., May, P. A., Kalberg, W. O., Kodituwakku, P., Gossage, J. P., Trujillo, P. M., et al. (2005). A practical clinical approach to diagnosis of fetal alcohol spectrum disorders: Clarification of the 1996 institute of medicine criteria. Pediatrics, 115(1), 39–47. 10.1542/peds.2004-0259

41. Bower, C., Elliott, E. J., Zimmet, M., Doorey, J., Wilkins, A., Russell, V., et al. (2017). Australian guide to the diagnosis of foetal alcohol spectrum disorder: A summary. Journal of Paediatrics & Child Health, 53(10), 1021–1023. 10.1111/jpc.13625

42. Coles, C. D., Gailey, A. R., Mulle, J. G., Kable, J. A., Lynch, M. E., & Jones, K. L. (2016). A comparison among 5 methods for the clinical diagnosis of fetal alcohol spectrum disorders. Alcoholism, clinical and experimental research, 40(5), 1000–1009. 10.1111/acer.13032

43. Scottish Intercollegiate Guidelines Network (SIGN) (2019). Children and young people exposed prenatally to alcohol (SIGN 156): a national clinical guideline. SIGN, Edinburgh. https://www.sign.ac.uk/our-guidelines/children-and-young-people-exposed-prenatally-to-alcohol/. Accessed 25 April 2024.

44. Sanders, J. L., Breen, R. E. H., & Netelenbos, N. (2017). Comparing diagnostic classification of neurobehavioral disorder associated with prenatal alcohol exposure with the Canadian fetal alcohol spectrum disorder guidelines: a cohort study. CMAJ Open, 5(1), E178–E183. 10.9778/cmajo.20160137

45. Cook, J. L., Green, C. R., Lilley, C. M., Anderson, S. M., Baldwin, M. E., Chudley, A. E., et al. (2016). Fetal alcohol spectrum disorder: a guideline for diagnosis across the lifespan. CMAJ, 188(3), 191–197. 10.1503/cmaj.141593

46. Landgraf, M. N., Nothacker, M., & Heinen, F. (2013). Diagnosis of fetal alcohol syndrome (FAS): German guideline version 2013. European journal of paediatric neurology, 17(5), 437–446. 10.1016/j.ejpn.2013.03.008

47. Okulicz-Kozaryn, K., Maryniak, A., Borkowska, M., Smigiel, R., & Dylag, K. A. (2021). Diagnosis of fetal alcohol spectrum disorders (FASDs): Guidelines of interdisciplinary group of Polish professionals. Int J Environ Res Public Health, 18(14). 10.3390/ijerph18147526

48. Reid, N., Shanley, D. C., Logan, J., White, C., Liu, W., & Hawkins, E. (2022). International survey of specialist fetal alcohol spectrum disorder diagnostic clinics: comparison of diagnostic approach and considerations regarding the potential for unification. Int J Environ Res Public Health, 19(23), 10.3390/ijerph192315663

49. Webster, B. M., Carlisle, A. C. S., Livesey, A. C., Deeprose, L. R., Cook, P. A., & Mukherjee, R. A. S. (2023). Evaluating the difference in neuropsychological profiles of individuals with fasd based on the number of sentinel facial features: a service evaluation of the FASD UK National Clinic Database. Children, 10(2), 266, 10.3390/children10020266

50. Astley, S. J., & Clarren, S. K. (2001). Measuring the facial phenotype of individuals with prenatal alcohol exposure: correlations with brain dysfunction. Alcohol and alcoholism, 36(2), 147–159. 10.1093/alcalc/36.2.147

51. Lange, S., Rovet, J., Rehm, J., & Popova, S. (2017). Neurodevelopmental profile of fetal alcohol spectrum disorder: A systematic review. BMC Psychology, 5(1), 22. 10.1186/s40359-017-0191-2

52. Lange, S., Shield, K., Rehm, J., Anagnostou, E., & Popova, S. (2019). Fetal alcohol spectrum disorder: neurodevelopmentally and behaviorally indistinguishable from other neurodevelopmental disorders. BMC Psychiatry, 19(1), 322. 10.1186/s12888-019-2289-y

53. OECD (2024). Researchers (indicator). 10.1787/20ddfb0f-en. Accessed 25 April 2024.

54. OECD (2024). Gross domestic spending on R&D (indicator). 10.1787/d8b068b4-en. Accessed 25 April 2024.

55. Coons-Harding, K. D., Azulai, A., & McFarlane, A. (2019). State-of-the-art review of transition planning tools for youth with fetal alcohol spectrum disorder in Canada. Journal on Developmental Disabilities, 24(1), 81–98.

56. May, P. A., Hasken, J. M., de Vries, M. M., Marais, A.-S., Abdul-Rahman, O., Robinson, L. K., et al. (2024). Maternal risk factors for fetal alcohol spectrum disorders: Distal variables. Alcohol, Clinical and Experimental Research, 48(2), 319–344. 10.1111/acer.15246

57. Astley, S. J., Bailey, D., Talbot, C., & Clarren, S. K. (2000). Fetal alcohol syndrome (FAS) primary prevention through fas diagnosis: II. A comprehensive profile of 80 birth mothers of children with FAS. Alcohol Alcohol, 35(5), 509–519. 10.1093/alcalc/35.5.509

58. Bandura, A. (1986). Social foundations of thought and action: a social cognitive theory. Englewood Cliffs, NJ: Prentice-Hall, Inc.

59. Kvigne, V. L., Leondardson, G. R., & Welty, T. K. (2006). Characteristics of fathers who have children with fetal alcohol syndrome or incomplete fetal alcohol syndrome. S D Med, 59(8), 337–340. 10.1016/j.jpeds.2004.07.015

60. Kvigne, V. L., Leonardson, G. R., Borzelleca, J., & Welty, T. K. (2008). Characteristics of grandmothers who have grandchildren with fetal alcohol syndrome or incomplete fetal alcohol syndrome. Maternal and Child Health Journal, 12(6), 760–765. 10.1007/s10995-007-0308-y

61. Kautz-Turnbull, C., Adams, T. R., & Petrenko, C. L. M. (2022). The strengths and positive influences of children with fetal alcohol spectrum disorders. American journal on intellectual and developmental disabilities, 127(5), 355–368. 10.1352/1944-7558-127.5.355

62. Kapasi, A., Makela, M. L., Flannigan, K., Joly, V., & Pei, J. R. (2019). Understanding employment success in adults with Fetal Alcohol Spectrum Disorder. Journal of Vocational Rehabilitation, 51(3), 377–393. 10.3233/JVR-191053

63. Currie, B. A., Hoy, J., Legge, L., Temple, V. K., & Tahir, M. (2016). Adults with fetal alcohol spectrum disorder: factors associated with positive outcomes and contact with the criminal justice system. Journal of population therapeutics and clinical pharmacology = Journal de la therapeutique des populations et de la pharmacologie clinique, 23(1), e37–52.

64. Corrigan, P. W., Lara, J. L., Shah, B. B., Mitchell, K. T., Simmes, D., & Jones, K. L. (2017). The public stigma of birth mothers of children with fetal alcohol spectrum disorders. Alcohol Clin Exp Res, 41(6), 1166–1173, 10.1111/acer.13381

65. Munn, Z., Pollock, D., Khalil, H., Alexander, L., McLnerney, P., Godfrey, C. M., et al. (2022). What are scoping reviews? Providing a formal definition of scoping reviews as a type of evidence synthesis. JBI Evidence Synthesis, 20(4). 10.11124/JBIES-21-00483.

66. Lasserson TJ, T. J., Higgins JPT. (2023). Chapter 1: Starting a review. http://www.training.cochrane.org/handbook. Accessed 19 April 2024.

67. Chasnoff, I. J., Wells, A. M., & King, L. (2015). Misdiagnosis and missed diagnoses in foster and adopted children with prenatal alcohol exposure. Pediatrics, 135(2), 264–270, 10.1542/peds.2014-2171

68. Geier, D. A., & Geier, M. R. (2022). Fetal alcohol syndrome and the risk of neurodevelopmental disorders: A longitudinal cohort study. Brain & development, 44(10), 706–714. 10.1016/j.braindev.2022.08.002

