## Supplementary information 1_search strategies and supplementary figure for "Trends in fetal alcohol spectrum disorder (FASD) research: a systematic scoping review and bibliometric analysis"

### Online Resource 1
### Supplementary information for: Trends in fetal alcohol spectrum disorder (FASD) research: systematic scoping review and bibliometric analysis

Dr Cheryl McQuire[1] Nessie Felicia Frennessen, James Parsonage, Molly Van der Heiden, Dr David Troy, Dr Luisa Zuccolo

[1] Corresponding author Dr Cheryl McQuire, University of Bristol, UK:, Bristol Medical School, Centre for Public Health, University of Bristol, Canynge Hall, 39 Whatley Road, Bristol, UK, BS8 2PS

Supplementary Table S1: Search Strategy for OVID Medline

| # ▲ | Searches | Results |
|---|---|---|
| 1 | Fetal Alcohol Spectrum Disorders/ | 4612 |
| 2 | f?etal alcohol syndrome.tw. | 2456 |
| 3 | FASD.tw. | 1561 |
| 4 | Fetal alcohol spectrum disorder.mp. | 1098 |
| 5 | Foetal alcohol spectrum disorder.mp. | 44 |
| 6 | Alcohol Related Neurodevelopmental Disorder.mp. [mp=title, book title, abstract, original title, name of substance word, subject heading word, floating sub-heading word, keyword heading word, organism supplementary concept word, protocol supplementary concept word, rare disease supplementary concept word, unique identifier, synonyms] | 99 |
| 7 | ARND.mp. [mp=title, book title, abstract, original title, name of substance word, subject heading word, floating sub-heading word, keyword heading word, organism supplementary concept word, protocol supplementary concept word, rare disease supplementary concept word, unique identifier, synonyms] | 101 |
| 8 | 1 or 2 or 3 or 4 or 5 or 6 or 7 | 5953 |
| 9 | limit 8 to yr="2000 -Current" | 3721 |

Supplemental Figure S2: Number of articles published per year for thematic groupings with > 50 published articles between 2000 – 2023

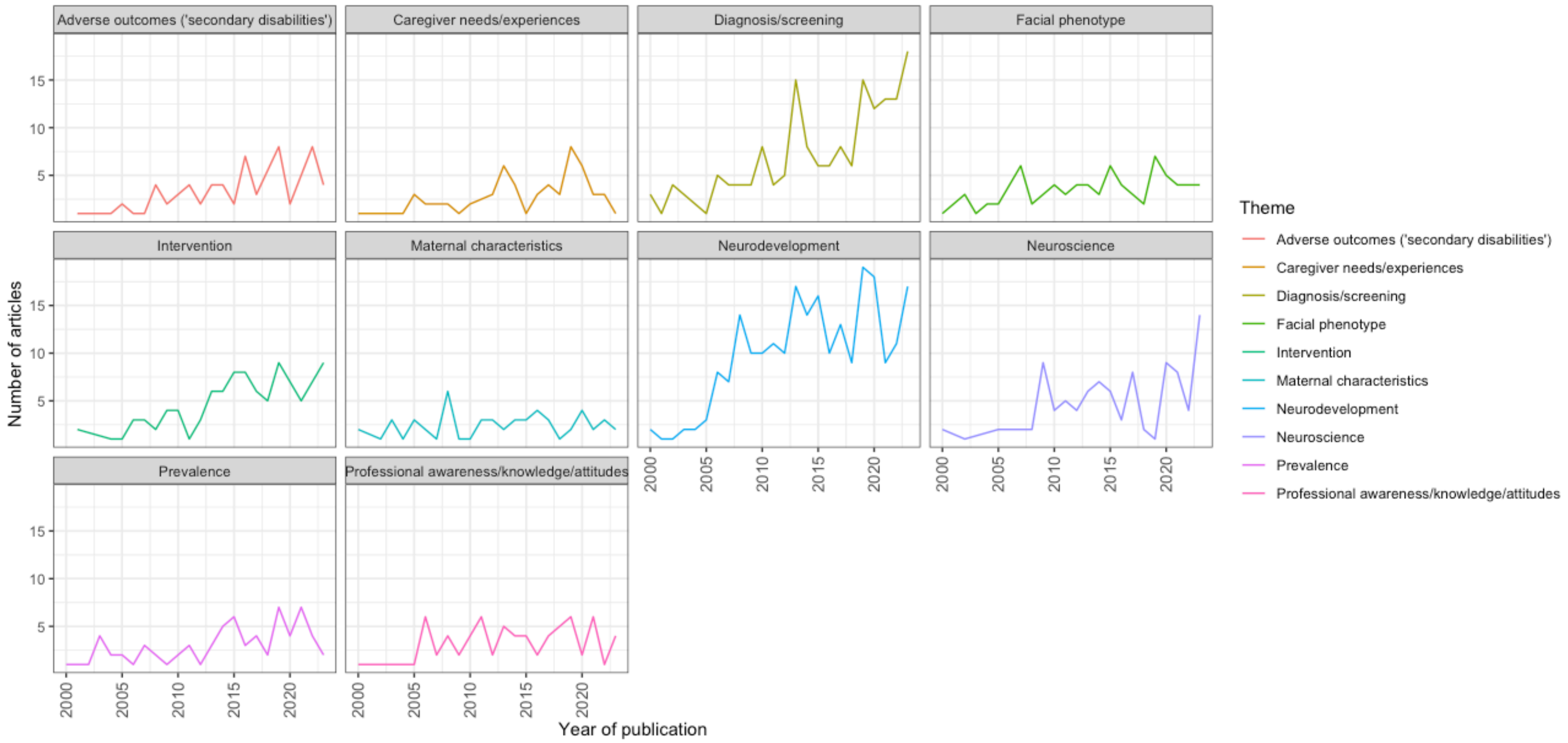

Supplementary Table S3: Search Strategy for comparative review of FASD, ADHD and autism in PubMed

| **Limits 2000/1/1 - 2023/12/31, humans, search terms in title** | |
|---|---|
| **Date search conducted: 08.04.24** | |
| **Name of neurodevelopmental condition** | **Search terms used** |
| FASD | (((((("fetal alcohol spectrum disorder"[Title]) OR (foetal alcohol spectrum disorder[Title])) OR ("FASD"[Title])) OR ("fetal alcohol syndrome"[Title])) OR ("foetal alcohol syndrome"[Title]) ) OR ("Alcohol Related Neurodevelopmental Disorder"[Title])) OR ("ARND"[Title]) |
| ADHD | ("ADHD"[Title]) OR ("attention deficit hyperactivity disorder"[Title]) |
| Autism | ("autism"[Title]) OR ("autistic"[Title]) |
